# Patterns of Suicidal Stress Disclosure on Social Media: Integrating Computational and Qualitative Approaches

**DOI:** 10.1101/2025.06.09.25329115

**Authors:** Hayoung Kim Donnelly, V. Scott H. Solberg, Jennifer G. Green, Derry Wijaya, David A. Jobes

## Abstract

The lack of understanding of how individuals communicate suicidal stress hinders global suicide intervention plans and practices. This study identifies patterns of online suicidal disclosure by applying natural language processing and thematic analysis to 128,587 narratives written by individuals at risk of suicide on an international social media platform between 2021 and 2022. Six unique themes emerged, reflecting heterogeneous behavior patterns: repetitive suicidal ideation disclosure (36.4%), relational stress (31.9%), suicide attempts and negative healthcare experiences (9.9%), abuse (8.8%), contextual stress (7.2%), and philosophical or informational discussion (5.8%). Across these clusters, three common patterns also emerged-social disconnection, manifestations of stress, and feelings of burdensomeness-highlighting shared underlying experiences. Integrating psychological theory and clinical guidance, this study introduces a typological framework that links data-driven insights with online behavioral patterns related to suicide. Our findings contribute to informing global suicide prevention strategies by providing real-life examples of diverse suicidal expressions.

## INTRODUCTION

### Health Disparities and Suicidal Disclosure

Suicide has been a global health concern for decades, with more than 700,000 people lost by suicide every year (World Health Organization, 2021). The disparities between relatively more and less privileged communities make this situation even more alarming. For instance, more than 70% of global suicide occurrences are in low and middle-income countries (World Health Organization, 2021). The disparities persist within individual countries. In Ethiopia, people who experienced homelessness and financial crisis reported higher suicide risk than others (Yohannes et al., 2023). In the U.S., marginalized populations, including sexual and gender minorities and incarcerated individuals, reported a higher suicide rate than others (Centers for Disease Control and Prevention, 2022). North Korean Refugees in South Korea also recorded a higher suicide risk than other citizens in South Korea (Nam et al., 2021). These significant disparities within and across countries underscore the urgent need for global mental health equity initiatives.

Among scholars and researchers focused on suicide, the phenomenon of low disclosure among people with suicidality has been a particular concern in preventing suicide (An et al., 2023; Daudi et al., 2023; Hom et al., 2015; Eskin et al., 2015; Jordans et al., 2018; Hagaman et al., 2017; Zewdu et al., 2019). More than half of people with suicide risk report never sharing their challenges with others (Hom et al., 2015; Husky et al., 2016; Rickwood & Thomas, 2012), and this low disclosure rate is commonly observed across countries (An et al., 2023; Eskin et al., 2015; Jordans et al., 2018; Hagaman et al., 2017; Zewdu et al., 2019). The internal and public stigma, fear of involuntary interventions, and lack of social support are considered common barriers to suicidal disclosure. Marginalized populations (e.g., those defined by racial or sexual identities, disability status, immigrant status, or socioeconomic status) experience additional stigma, such as identity-related stigma, in addition to mental health-related stigma (Hom et al., 2015; Nam et al., 2021; Pretorius et al., 2019). The additional barriers faced by marginalized individuals may exacerbate disparities in suicide between communities.

So far, knowledge on suicidal disclosure has been primarily shaped by non-naturalistic contexts and with structured data, such as administrative or surveys in medical systems. A limitation of the current knowledge is that it may not fully capture how individuals naturally describe their suicidal experiences in daily life, particularly among diverse populations who do not have access to medical settings. Therefore, innovative strategies could help build more inclusive knowledge and initiatives by capturing diverse suicidal disclosure types from broader communities. Such strategies can involve collecting and analyzing (1) data generated naturally in people’s daily lives rather than data collected from clinical or research settings (Ophir et al., 2020), (2) narrative data rather than quantitative data (Jobes et al., 2004), and (3) data that includes stories from non-traditional samples such as social media posts (Chancellor & De Choudhury, 2020).

### Online Suicidal Disclosure

The current study focuses on online narratives of suicidal disclosure. Online spaces have become a common platform for people to share health concerns and seek health-related information, especially following the COVID-19 pandemic (Ellis et al., 2021; Inthiran, 2017; Pretorius et al., 2019; Thornton et al., 2017; Wilks et al., 2019). People report using the internet to find information (Datareportal, 2021), almost three out of four internet users in the U.S. have searched for health information online within a year (Fox & Duggan, 2013). Compared to face-to-face disclosure, researchers have consistently observed that individuals feel more comfortable disclosing their sensitive and personal problems online (Haner & Pepler, 2016). The preference for online disclosure is also associated with the inherent strengths of the online space, such as high accessibility to communities (i.e., no time and space restrictions) and low risk of stigma from anonymous communication (Frost et al., 2016; Rose & Friedman, 2013). Like face-to-face help-seeking behavior, online help-seeking behavior has been found to promote healthier behaviors and enhance the effectiveness of healthcare services (Tan et al., 2017).

In addition to lowering stigma and increasing accessibility, particularly for mental health-related disclosure, individuals may perceive that online spaces allow them to be autonomous and to easily connect with others who share similar experiences (Bell et al., 2018). First, people want to be self-determinant, self-reliant, and autonomous to decide their next step, including whether to take the advice of others after disclosures (Sheehan et al., 2019). When receiving advice online, people feel less obligated to take that advice and to put that advice into action (Pretorius et al., 2019). People worry less about unwanted interventions, such as involuntary hospitalization and treatment, when disclosing online versus offline spaces (van der Schyff et al., 2023). Even with this autonomy, people who disclose their suicidality online are more likely to use professional mental health services than those who do not (Frost & Casey, 2016). Second, people find that it is easier to meet others who have similar experiences with them in online spaces (Bell et al., 2018). Finding others with similar experiences online makes people aware that they are not the only ones struggling with suicidal issues (Bell et al., 2018). This has been shown to lead to positive outcomes after disclosing suicidality online, such as inducing feelings of relief and acceptance (Bell et al., 2018).

### Limitations in Previous Studies

Although an increasing number of studies actively explore online suicidal disclosure, a major concern is the low explainability and interpretability of their findings. Previous research has primarily focused on predicting the level of suicide risk of individuals by developing accurate and efficient machine-learning models (Chancellor & De Choudhury, 2020). This line of work demonstrates the potential of timely, cost-effective risk detection systems with high accuracy and sensitivity in prediction. However, a limitation of previous research is the low interpretability of findings, which often fail to illustrate the unique contexts of individuals of the complex mechanisms of suicidal ideation and behaviors (Zhang et al., 2022). This is especially a barrier to translating their findings into actionable suicide prevention efforts, as they may be difficult for clinicians and policymakers to understand and apply.

Another limitation is the lack of evidence derived from people’s narratives, as opposed to quantitative features of online behaviors. For instance, quantitative features (e.g., number of posts or comments) and linguistic features (e.g., forms of nouns, verbs, and pronouns) are commonly used to understand online suicide-related behavior (De Choudhury & De, 2014; Homan et al., 2022; Monselise & Yang, 2022). The advantage of these metrics is that they can provide suggestions for online suicide prevention strategies, such as sharing resources on platforms beyond suicide-focused ones based on these users’ activity-focused behavior (Monselise & Yang, 2022). However, understanding what and how people actually talk about their suicide risk is crucial for developing contextualized and individualized online interventions.

### The Current Study

The major research questions are “What are the different types of suicide disclosures observed on social media platforms?” To answer this question, this study utilizes suicidal disclosure narratives from a social media platform, specifically a Reddit sub-community called *r/SuicideWatch*, where individuals voluntarily write in an anonymous setting. A mixed-method approach, combining Natural Language Processing and Qualitative Thematic Analysis, was developed and applied to data from r/SuicideWatch with the goal of offering interpretable and accessible insights into suicidal disclosure from a large dataset. Further, the identified types from narratives are explained using existing suicide theories, and potential intervention strategies for each disclosure type are discussed.

## METHOD

### Data

A total of 128,587 posts from r/SuicideWatch, spanning January 1st, 2021, to December 31st, 2022, were collected by the Brandwatch web crawler using the Reddit Application Programming Interface (API). r/SuicideWatch is a subreddit dedicated to “peer support for anyone struggling with suicidal thoughts,” where users can discuss any suicide-related topics. Based on Reddit Statistics in 2022, over 70% of users are from countries using English as a primary language, 68% of Reddit users are male, and 51% of Reddit users are between the ages of 18 and 29 (Reddit Statistics, 2022). In this study, each r/SuicideWatch post and username did not include identifiable information, therefore, this research qualifies for an exemption from the Institutional Review Board process at [the authors’ institution]. The posts were written by 76,547 users, with a median of one post per user. Among the different features of Reddit, this research specifically focused on analyzing the full text of posts from the dataset (Figure 1). The data and analysis code used in this study are available strictly upon request to the authors.

**Figure 1.**
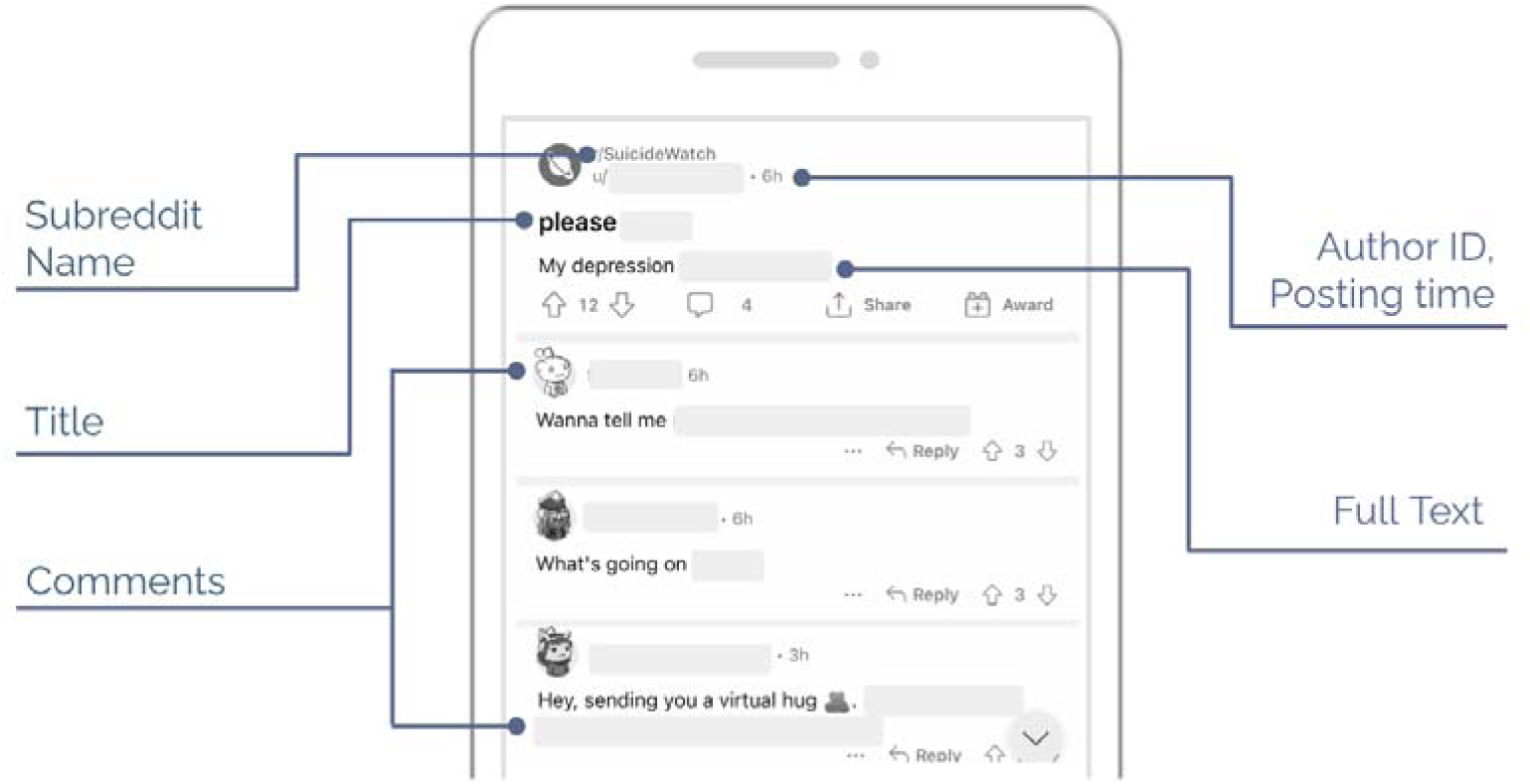
Reddit features

### Analytical Approach

This study developed and employed a mixed-methods approach, combining natural language processing (NLP) with qualitative thematic analysis (Figure 2). The usefulness of NLP has been highlighted due to its ability to automatically analyze diverse types of data, including social media posts, health records, and interview transcripts (Zhang et al., 2022). Specifically, this study employed Latent Dirichlet Allocation (LDA), a widely used unsupervised learning technique. LDA organizes numerous documents into a limited number of topic clusters and identifies keywords and key documents that best represent each cluster. This study compared the perplexity scores of different LDA models with varying numbers of clusters. A model with the lowest perplexity was selected as the best classification model (Blei et al., 2003; Williams et al., 2020).

**Figure 2.**
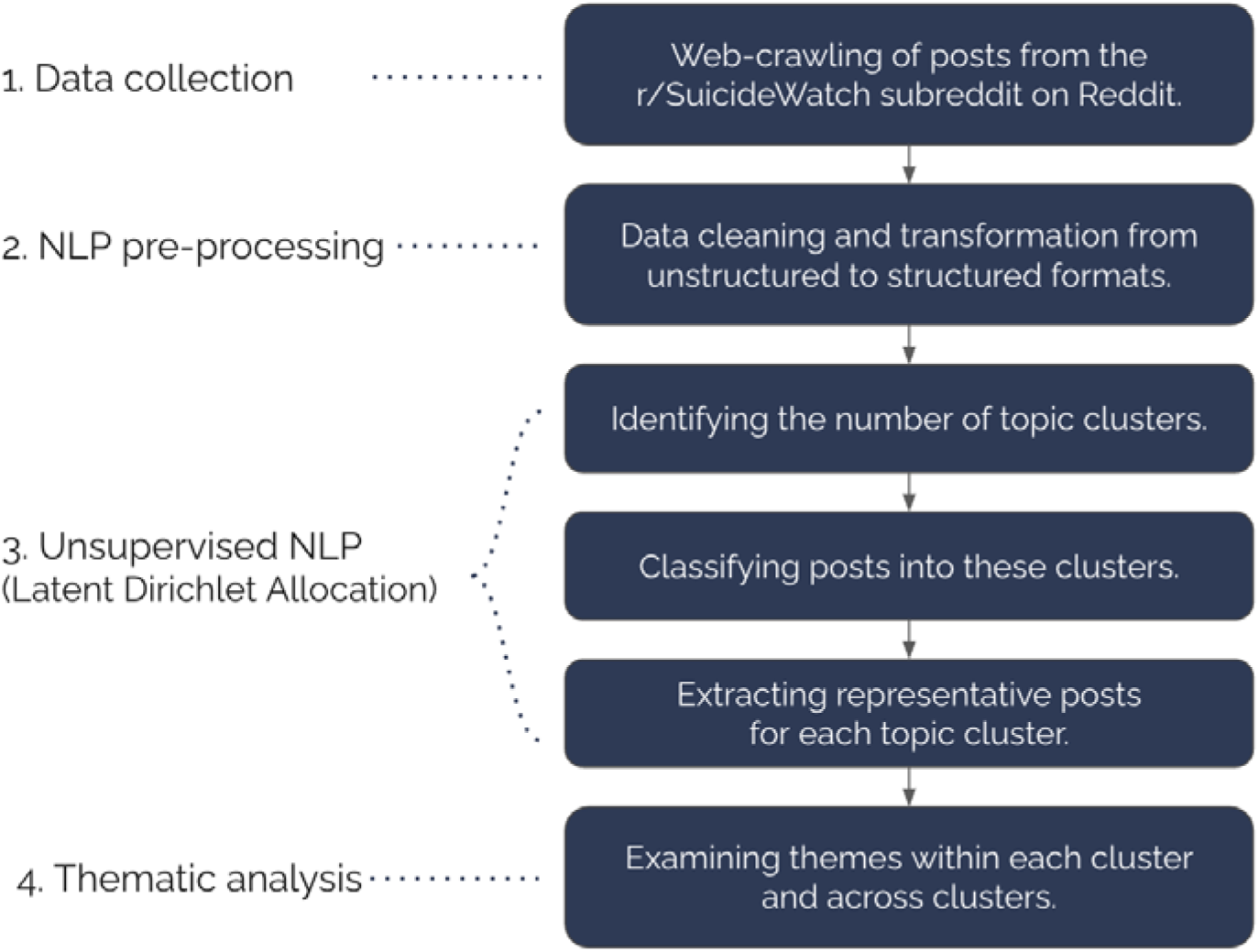
Research Process

Although LDA has strengths in providing statistical evidence for the number of clusters while summarizing numerous documents, it lacks a systematic approach for interpreting the latent topics within the identified clusters. To address this, the study integrates qualitative thematic analysis to provide more detailed and interpretable insights. Five professionals, including two doctoral candidates, two graduate students, and one scientist in social science, analyzed the posts using an open coding process to identify themes. Given the extensive data volume (128,587 posts), analyzing all posts manually was not feasible. Therefore, the top 30 key documents for each cluster identified from the LDA were only evaluated by coders. The coding results were cross-validated, and only those with agreement from more than three coders were included as final.

## RESULTS

### Number of Latent Topic Clusters

The initial grid search recommended an LDA model with three latent topic groups as the best fit with the lowest flexibility (Figure 3). However, summarizing massive data and diverse types of suicidal disclosures into only three categories might lead to overlooking valuable information. To address this concern, an additional LDA grid search was performed. Three sub-datasets derived from the initial grid search were individually developed into their own LDA models. As a result, two topics were recommended as the optimal number for each sub-dataset with the lowest flexibility (947.49 in the first sub-dataset, 1508.8 in the second sub-dataset, and 1264.8 in the third sub-dataset). In other words, this hierarchical LDA grid search identified six topics as the optimal model for categorizing 128,587 posts.

**Figure 3.**
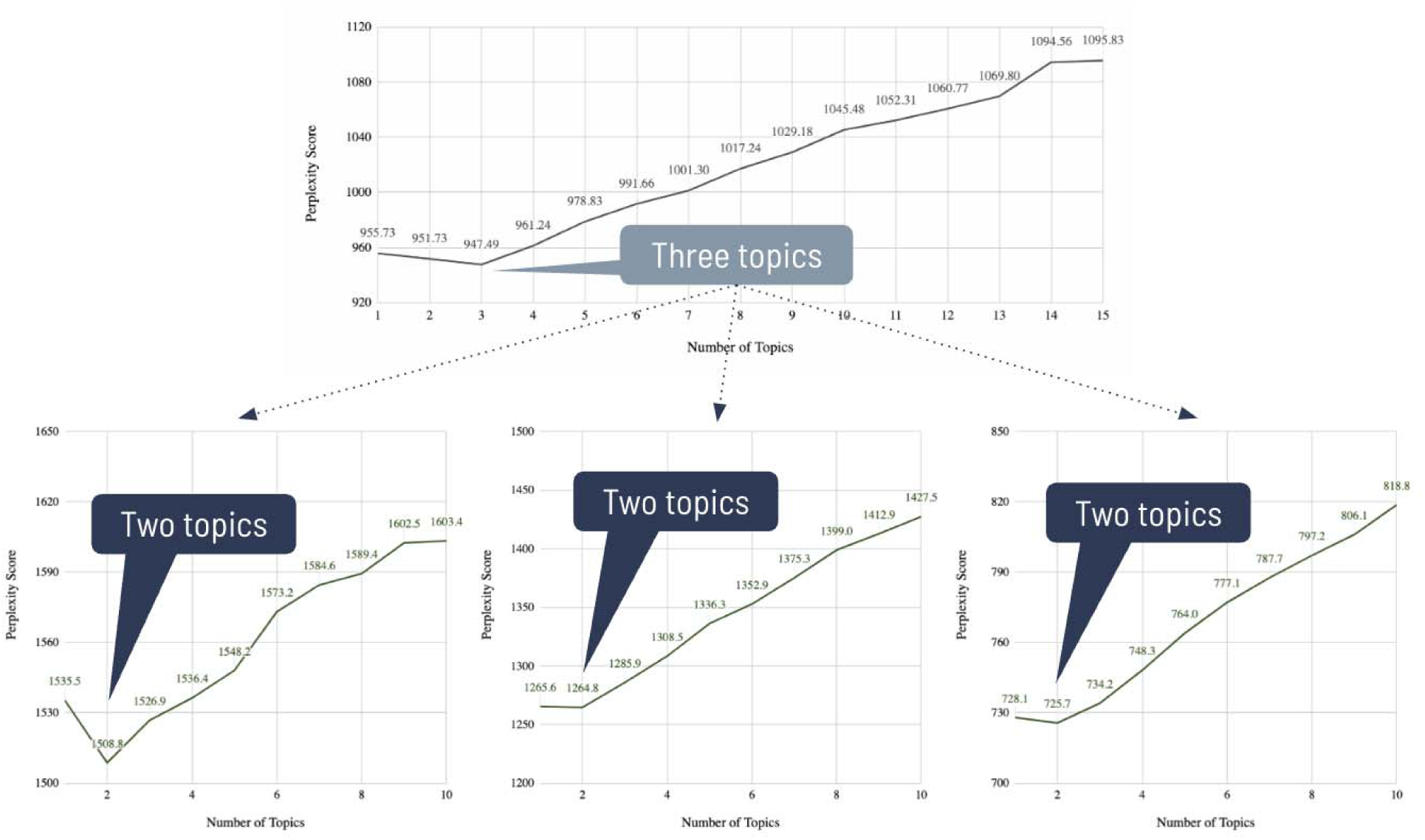
Hierarchical LDA grid search

### Unique Themes

The themes of the six clusters include (1) Repetitive suicide ideation, (2) Relational stress, (3) Suicide attempts and healthcare experiences, (4) Abuse, (5) Contextual stress, and (6) Philosophical and Informative Discussion (Figure 4).

**Figure 4.**
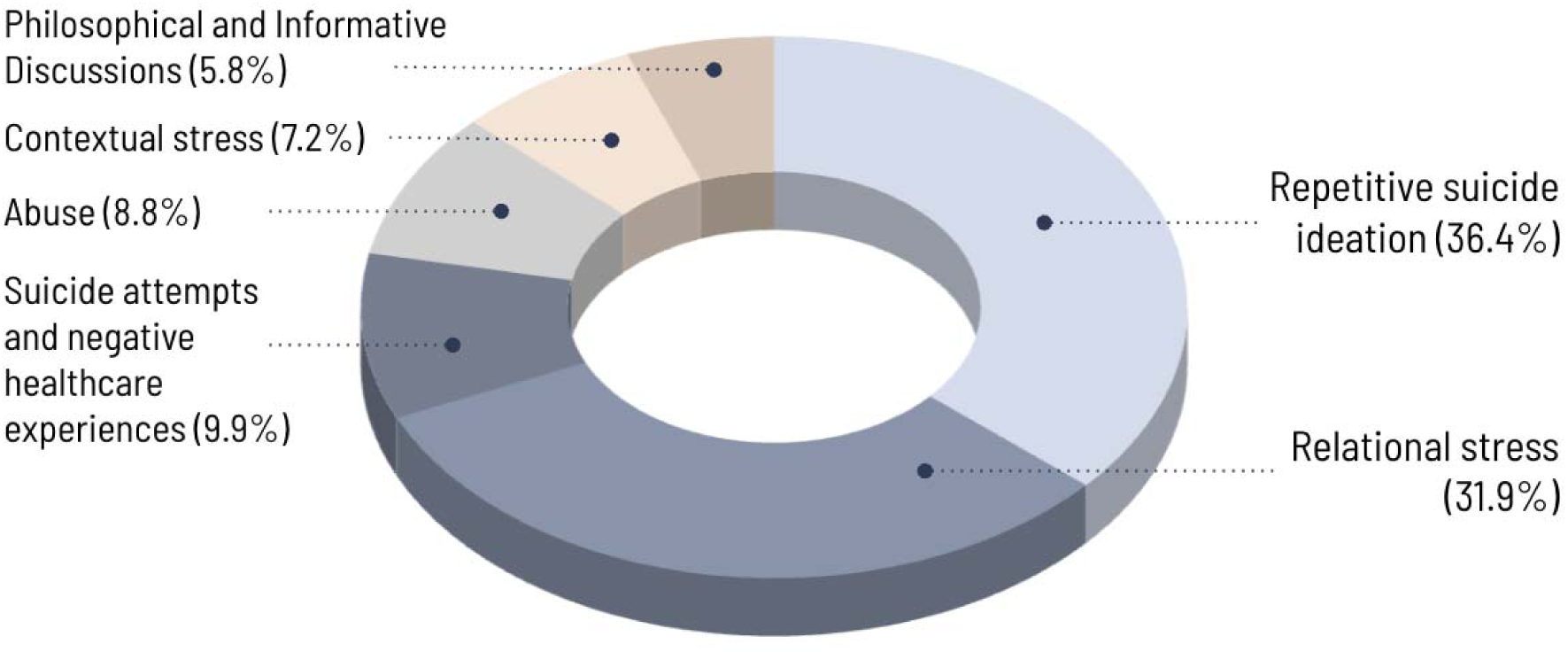
Themes and distributions of the six clusters

#### Theme 1: Repetitive suicide ideation

The first topic was identified as 36.4% of posts (n = 46,815) and the most common theme across the six clusters. People disclosed their thoughts, wishes, and intentions related to suicide over and over by using short, simple, and first-person-centered sentences (I-sentences). People’s disclosure is disproportionally focused on describing acute suicidal experiences at the moment of writing posts rather than describing past suicide-related experiences. For instance, people declared their wish to die (i.e., *“I wanna die.”, “I want to fucking die.”*), intention to kill themselves (i.e., *“I’m going to kill myself”*), and self-loathing (i.e., *“I’m useless”, “I want to punish myself. I deserve it.”*) in their posts. Especially, there was a clear common pattern that they repeated these declarations many times (i.e., *“I wanna die. I wanna die. I wanna die.…*[repeated]*”, “I’m going to kill myself I’m going to kill myself I’m going to kill myself …* [repeated]*”*).

#### Theme 2: Relational stress

This topic was the second most common topic among the six topics and was identified as 31.9% (n = 41,049) of posts. People disclosed negative relationships with different family, friends, and romantic partners, and how those experiences are related to their suicidal risk. Family-related disclosure included arguments, conflicts, negative experiences with family members (i.e., *“it’s the way that [parent] treats me and how she shows 0 respect for me.”*, “*I basically had a bad relationship with my siblings*”). They also shared negative reactions of family toward their disclosure of suicidality (i.e., “*aren’t you the one who tried to kill yourself last year?”*). Friend-related disclosure included having no friends (i.e., “*I barely have any friends”*), fear of making friends (i.e., *“That pissed me off a bit because I can’t exactly stop a nightmare inducing fear to make friends.”*), and conflicts with friends (i.e., *“We had a couple of arguments and I don’t think we’re friends anymore.”*). Romantic relationship-related disclosure includes experience in breakup (i.e., “*My sweet, kind, wonderful college boyfriend who pretty much felt like my soulmate dumped me, and it was all my fault.”),* an affair of a romantic partner (i.e., *“after my ex cheated on me (after 3 years) i was a complete wreck and attempted”*), and low confidence in dating (i.e., *“I’d love to have a boyfriend, but I know that won’t happen, probably ever because first of all, who’d want me”*). Additionally, some stress was caused by interaction between different social relationships. For instance, people disclosed relationship stress from negative experiences between family and romantic partners (i.e., *“Me and my parent and girlfriend had a huge fight. My parent hates me, my gf doesn’t want to see me and my family again. My fight with my parents lasted for 1 year.”*).

Additionally, in this cluster, people often disclosed they are either adolescents (i.e., *“today I turned [age]”*) or emerging adults (i.e., *“Some background, I’m a [grade] college student”).* People here also asked advice on how to support their significant others with suicide risk (i.e., *“I don’t know what to say to help him, or even how to act when I see him in person again this Friday. I feel so bad for him, I just want to help. Any advice?”*).

#### Theme 3: Suicide attempts and negative healthcare experiences

This topic was identified in 9.9% (n = 12,729) of posts and is the third most common topic among the six topics. People disclosed their past suicide attempts and asked for advice on means of self-harm in their posts (i.e., *“I tried suicide twice but got scared but the thoughts have always been around.”*; *“Is putting 200 metformin tablets in a blender, then mixing the fragments with alcohol a good way to go?”*). People described the onset of suicidality (i.e., *“From age [age], I’ve been struggling with suicidal thoughts”*) as well as the long history with suicidality (i.e., *“I am now [age] and have battled since my earliest years.”*). People shared their history of other mental health challenges (e.g., *“I was diagnosed with unipolar depression.”*).

People described their negative experiences with mental health services. They complained about negative interactions with professionals (i.e., *“I had a hard time being in the psych ward. The nurses ignored me.*”), a long wait time (i.e., *“I currently don’t have therapy since I’m on 2 different waiting lists for a new one.”*), and side effects from medicine (i.e., *“The medicine makes me drowsy. Recently, my anxiety seems to have turned to depression.”*). People felt tired and hopeless after trying different mental health treatments and perceived that these treatments did not help reduce their suicidality (*“Nothing is helpful, not even therapy.”, “More pills, therapy, talking, all the works. Didn’t work.”*). When they experienced no positive impact from treatment, people reported feeling tired, defeated, and lost (*“Enough is enough. I have tried so many different treatments, therapies, and drugs over the years.”, “Everything is such a failure. The psychiatrists. The psychologists. The counselling that goes nowhere.”*, *“What do I do? I just feel lost and no clue what to do anymore.”*).

#### Theme 4: Abuse

The 8.8% (n = 11,295) of posts were about anecdotes of abuse victimization with their suicidal risk. People disclosed their experience with verbal abuse (i.e., “*You don’t deserve to be my [child]” “My [siblings] started to disrespect me, started to call me names mainly “Fag, Gay, HOMO” I hated it.”*). People disclosed their experience with physical abuse (i.e., *“Proceeded to hit me with the plank. After this I got dragged to my room and got beaten again.”*). People disclosed their experience with sexual abuse (i.e., *“When I was [age], I was molested by my cousins… Suddenly she tug my shirt off and my pants off. I was frozen, I didn’t know what to do, I could not move”*). This cluster tended to focus on only one or a few specific incidents of negative social experiences, while the Disclosure of Relational Stress cluster described multiple examples of social experiences in daily situations. Additionally, people here often described their abuse experience happening in their early childhood (*“I’m like, “Stop it,” COMING FROM a [age]”, “We never wanted to know, a little [age] not knowing how to say or ask for help”, “When I was [age] I was molested by my cousins.”, “from the age of [age] I was mentally and physically abused by him”*).

#### Theme 5: Contextual stress

The 7.2% (n = 9,254) of posts disclosed stress from the context of living related to finance, imprisonment, the COVID-19 pandemic, and career. People disclosed their current financial stress (i.e., *“every moment was a living fear of the debt collector knocking on the door”*), absence of insurance (*“I had no insurance”*), and housing instability (i.e., *“My landlord said I would be able to pay off my portion of the rent gradually with part time jobs while he looked for replacement tenants.”*). People also shared their or their significant others’ imprisonment history (*“I went to 3 prisons for a year and a half and I sat in jail for about 3 years.”*; *“my [parent] got sent to jail again shortly”*), while they disclose their suicidal stress. People mentioned the COVID-19 pandemic and its impacts on their daily life, such as social isolation due to the pandemic (i.e., *“I was able to study but with Covid I was left alone with my thoughts and kept thinking about what would happen if I failed again”*). The career-related issues include losing or fear of losing a job (i.e., *“I lost my job due to cost cutting measures.”),* stress in searching for a job (i.e., *“I have been job hunting and interviewing for weeks and gotten mostly rejections or just gotten ignored for the most part.”*), as well as academic stress (i.e., *“I started losing focus on my study”*; “*my grades were excellent but I was still disappointed”*).

#### Theme 6: Philosophical and informative discussion

The 5.8% (n = 7,445) of posts were about people’s thoughts and information about suicide, life, death, and mental health, rather than disclosing their own suicidal stress. This includes philosophical and metaphorical discussions (i.e., *“After death, the physical begins to deteriorate and life/energy is simply moved to another being.”*) and sharing information related to mental health (*“Depression differs in type, - a common definition of depression is ‘sub-optimal psychological function’, …”*).

### Common Themes

While identifying each cluster’s unique themes, three common patterns across the clusters were also found. First, people commonly said that they don’t have anyone to talk to about their suicidal issues (i.e., *“I have no one to talk to”, “I don’t know who else I can talk to or rely on.”*). Second, people often said that they wrote posts because they just wanted to talk to someone about their suicidal stress (i.e., *“I’m using this post as a place to vent as I feel maybe someone can help or make me feel better”, “I guess I just wanted to make this post because I feel like I need to vent”*). They also expressed their appreciation for letting them share their suicidal risk (i.e., *“Thank you Reddit for letting me get this off my chest safely.”).* Lastly, people often apologized to others who would read their posts (i.e., *“I’m sorry this was so long and I hope don’t come off as too whiny.”*). These themes were observed in all clusters except the Philosophical and Informative Discussion cluster.

## DISCUSSION

People express their struggles with suicidal risk in unique ways on social media, using their own language. The findings of this study reveal the distinctive types of suicidal disclosure as well as the common psychological characteristics across these disclosures. The current study discusses each disclosure type with the existing suicide theories and proposes different intervention strategies tailored to each type by adopting the approach of Collaborative Assessment and Management of Suicidality (CAMS). CAMS, which is an evidence-based, suicide-focused framework, identifies diverse suicidal drivers from patient narratives such as relationship, psychological distress, and vocational concerns (Lynch et al., 2024). It is an effective and flexible approach that emphasizes collaborative intervention design, incorporating existing suicide prevention strategies that best fit the individual’s context.

### Suicidal Disclosure Type 1: Repetitive Suicide Ideation

In the U.S., 5 out of every 100 adults (18 and older) and 12 out of every 100 young adults (18 to 25) reported experiencing suicidal ideation in the past year (SAMHSA, 2023). People experience unbearable psychological pain from repetitive suicidal thoughts and feel powerless when they cannot stop them (Kerkhof & van Spijker, 2011). Greater attention to suicidal ideation, alongside suicide attempts and deaths, is essential for supporting individuals at risk (Jobes et al., 2024). This study found that more than one-third of r/SuicideWatch posts expressed repetitive suicidal ideation by duplicating the same sentence in a single post. Individuals may repeat the same sentence of suicidal ideation multiple times by either manually writing it repeatedly or copying and pasting it. Either way, it suggests that these thoughts are not fleeting or transient; and that they may not dissipate quickly.

Repetitive suicidal thinking increases the intensity of suicide ideation (Law & Tucker, 2018; Rogers & Joiner, 2018), which in turn elevates the risk of completed suicide and suicide attempts (Rossom et al., 2017). People in this cluster often wrote posts with present tenses and simple verbiage, which may have reflected their acute and imminent suicide risk. Prior studies have shown that more than half of individuals attempted suicide within one hour of experiencing suicidal ideation (Deisenhammer et al., 2009), and 70% attempted suicide within one hour of forming a plan (Simon et al., 2001). Therefore, time-sensitive intervention for acute suicidal risk is crucial to prevent the progression from suicidal ideation to an attempt (Gao et al., 2018; Renjith et al., 2022).

The Safety Planning Intervention (SPI; Stanley & Brown, 2012) and Crisis Response Planning (Bryan et al., 2017) can be an effective intervention for addressing repetitive and present suicidal thoughts. For example, SPI is a widely recognized suicide crisis intervention that aims to help individuals identify warning signs and develop coping strategies (Stanley et al., 2008). Notably, the step of identifying distractors in SPI helps individuals redirect their focus away from suicidal thoughts (Chesin et al., 2016; Stanley & Brown, 2012).

### Suicidal Disclosure Type 2: Relational Stress

The Relational stress category in this study may describe thwarted belongingness, as conceptualized in the Interpersonal Psychological Theory of Suicide (IPTS; Joiner et al., 2009; Van Orden et al., 2010). Prior studies have found that people who experienced negative relationships with their family members (i.e., parents, siblings, extended family) were more likely to be at high suicide risk (Mubashar & Butt, 2022). Negative peer relationships, including low levels of peer connectedness, bullying, and victimization, have also been related to the risk of suicidality (Cui et al., 2011). For example, evidence has shown that, not long before the onset of suicidal risk or death by suicide, many people experienced negative events in romantic relationships, such as break-ups and affairs (Kazan et al., 2016).

Relational stress was often reported by adolescents and young adults in this study. Although the impact of negative social relationships on suicide has been identified among all age groups, it may be especially potent among adolescents and young adults (Diamond et al., 2022; Gençöz & Or, 2006). During adolescence and young adulthood, social relationships expand from caregivers and family to include friends, teachers, and romantic partners. In fact, after accounting for other behavioral health such as depression, social relationships emerged as the most relevant factors of suicidal ideation among youth (Donnelly et al., 2023).

As one example of an intervention related to relational stress in the context of suicidality, Dialectical Behavior Therapy (DBT) includes training in interpersonal effectiveness skills, emotional and distress regulation skills, and mindfulness for individuals at suicide risk (Lenz et al., 2016). DBT has been suggested as a valuable psychosocial intervention program for youth with relational stress and suicidality (Glenn et al., 2019). Additionally, a few people in the current study expressed that they don’t know what to do to support their significant others with suicidal risk. This finding highlights the need to provide psychoeducation for the general population, such as suicide literacy education and gatekeeper training to help people support their significant others (Batterham et al., 2013; Shin & Kim, 2013).

### Suicidal Disclosure Type 3: Suicide Attempts and Negative Healthcare Experiences

Research has shown that past suicide attempts drive future suicide risk behaviors (Wexler et al., 2008). People with chronic suicidality reported higher risk of suicide attempts than those with short-term suicidal stress (Oquendo et al., 2006). People who have suicidality and other mental health reported a high risk of suicide compared to people who only have suicidality (Svanborg et al., 2008). In this disclosure group, people also shared the means of suicide they have used in past attempts or asked for advice on the potential means for future suicide attempts, including overdosing on medications, using firearms, and hanging themselves.

People have diverse experiences with mental health services, including using psychiatric medication, engaging in psychotherapy, visiting the emergency room, utilizing suicide hotline services, and undergoing hospitalization. However, people were often unsatisfied with mental health services due to a lack of empathy from professionals, repeated intake processes with different clinicians, and extended wait times for appointments. After experiencing no positive results from multiple therapy sessions and various medication trials, people often describe themselves as feeling lost and unsure of how to improve beyond these efforts. This symptom of learned helplessness (Maier & Seligman, 1976) is a critical barrier to intervening in suicidal risk (Wilson & Deane, 2010; Staiger et al., 2017). It minimizes hope, help-seeking, and continued engagement with the healthcare system (Wilson & Deane, 2010; Staiger et al., 2017).

These narratives underscore the need for systemic and institutional improvements in mental healthcare for high-risk groups. Clinicians should be sensitive to patients’ histories of suicidality and their experiences with mental health services (Shea, 1998). Increasing the number of mental health professionals is urgent to reduce waiting periods for individuals seeking psychotherapy or psychiatric services. Offering diverse mental health services beyond individual therapy and medication, such as community-based interventions and meditation classes, will provide individuals with opportunities to explore various interventions that may work for them.

### Suicidal Disclosure Type 4: Abuse

People who experienced sexual, physical, and verbal abuse were more likely to report suicidal thoughts, planning, and attempts than people who did not (Bruffaerts et al., 2010; Miller et al., 2013). Escape theory explains that people consider suicide to escape from the unbearable pain of traumatic memories (Shneidman, 1993). People who experienced abuse in early childhood reported a higher risk of suicide throughout their lifespan (Lopez-Castroman et al., 2013). However, self-hatred, self-blame, and negative self-image among survivors of abuse hinder the disclosure of other mental health issues (McElvaney et al., 2022). Especially when abuse experiences were major suicidal drivers, people with abuse experiences were less likely to disclose their suicidality (Collin-Vézina et al., 2021; McElvaney et al., 2022).

Suicide interventions with a deeper understanding and intervention for feelings of shame and a negative self-image are also recommended to help people with abuse experiences (McElvaney et al., 2022). For people who have abuse experience as major suicidal drivers, the Collaborative Assessment and Management of Suicidality (CAMS) suggested diverse suicide intervention strategies, including “exposure therapy, insight-oriented work, clinical hypnosis, EMDR therapy, cognitive processing therapy, and the like.” (Jobes, 2016 p. 110). Other research highlighted that people could be overwhelmed with strong emotions or triggered by painful memories before, while, and after disclosure of abuse (McElvaney et al., 2014). Therefore, it is especially salient for clinicians to explore people’s feelings, thoughts, and behavioral urges, especially after disclosure of abuse and suicidality.

### Suicidal Disclosure Type 5: Contextual Stress

People described various living challenges (e.g., housing issues, the COVID-19 pandemic) or a combination of these challenges on social media. During the pandemic, people lost their job (Kawohl & Nordt, 2020), people struggled with their financial situations (Crayne, 2020; Thakur & Jain, 2020), and students reported difficulty focusing (Aftab et al., 2021; Barzani & Jamil, 2021). Often these different contextual factors were combined rather than described as a single living challenge (Crayne, 2020; Elbogen et al., 2020; Madianos et al., 2014). Especially, the current study found that people discussed legal violations and imprisonment issues when talking about their suicidal stress. In fact, the suicide rate is higher among individuals with incarceration experience compared to the general U.S. population (Carson, 2021).

Moving forward, systemic and instrumental support is essential to reduce the suicide risk of people with contextual stressors. Protection legislation for employers and supportive policies for unemployed people can reduce suicide risk (Shand et al., 2022). Begun et al. (2016) suggest that providing mental health support for individuals released from correctional facilities can help prevent both suicide and re-incarceration. Providing contextually sensitive tele-therapy practices is helpful in reducing the suicidal risk of people with specific contextual stress during the COVID-19 pandemic (Jobes et al., 2020).

### Common Themes of Suicidal Disclosure

Across the suicidal disclosure types mentioned above, three common themes emerged from the posts: absence of someone to talk to, manifestation of suicidal stress, and expressions of apology. Previous findings show that people used online communities to find others to discuss their suicidal stress with as an alternative to offline communities (Brown et al., 2022; Pretorius et al., 2019). It is common for individuals with suicidality to feel sorry and consider themselves a burden, regardless of how others perceive them (Van Orden et al., 2010). It is important to note that feelings of guilt, such as apologizing for lengthy posts or grammatical errors, observed in this study should not be easily interpreted as causes of suicidal risk. However, burdensomeness may be a common psychological characteristic among individuals with suicidality, regardless of whether it serves as a direct cause of suicidal risk (Van Orden et al., 2010).

### Additional Theme: Philosophical and Informative Discussions

In addition to the above themes, another latent theme—Philosophical and Informative Discussion—was also identified in the current study. r/SuicideWatch is primarily designed as a peer support group for individuals experiencing suicidal stress. However, some individuals may perceive this community as an open forum for philosophical or political discussions about suicide, given that it is the largest group that appears when searching for “suicide” on Reddit. Therefore, it is important to be careful when determining these individuals’ suicide risk. They may discuss this theme either due to personal suicide risk or out of intellectual curiosity about the topic.

### Limitations

Despite the strengths of this study, several limitations should be considered. Firstly, this research aimed to understand people’s experiences at a behavioral level by focusing on individual posts rather than each person. Analyzing multiple posts from individuals may reveal suicidal disclosure patterns at the individual level, helping in the understanding of comprehensive and longitudinal experiences. Secondly, the study focused on intrapersonal disclosures through social media posts but did not consider interpersonal interactions such as comments to the disclosures. Reactions and subsequent communications can influence future disclosures. Including both posts and comments to explore their relationships will be beneficial for understanding disclosure types more in-depth. Fourth, the final number of themes was intentionally limited to align with the number of clusters determined by LDA, ensuring a balance between the computational and qualitative approaches. Each theme cluster contained different sub-themes, and to gain a deeper understanding of online suicide disclosure types, future research is recommended to explore each type more in-details. Fifth, it is important to note that this study sample is limited to individuals who are actively engaged in social media, rather than representing the overall population. However, a promising trend has been observed, with significant increases in internet usage among underprivileged populations who are typically underrepresented in online data during and after the COVID-19 pandemic (Datareportal, 2021). Nevertheless, it is still recommended to interpret and apply the current research findings with caution when considering broader public suicide prevention efforts beyond the online space.

## Conclusion

How and what people disclose regarding their suicide-related stress is not uniform. Counselors and clinicians need to familiarize themselves with different disclosure types and be prepared to develop treatment plans that are sensitive to each individual’s unique narrative. The strength of this research lies in its use of alternative data and methods to propose a framework for understanding suicidal disclosure types in online spaces. Researchers and policymakers should continue to prioritize listening to diverse voices through inclusive data and interdisciplinary methodologies to strengthen public suicide prevention efforts.

## Data Availability

All data produced in the present study are available upon reasonable request to the authors.

